# *TSC1* mRNA processing alterations in tuberous sclerosis complex (TSC) patients with negative or inconclusive genetic testing results

**DOI:** 10.1101/2025.01.30.25321206

**Authors:** Arthur Bandeira de Mello Garcia, Taís da Silveira Fischer, Maria Clara de Freitas Pinho, Lucas Fernandesw Jataí, Patrícia Santos da Silva, Tiago Finger Andreis, Larissa Brussa Reis, Cristina Brinckmann Oliveira Netto, Patricia Ashton-Prolla, Clévia Rosset

## Abstract

Tuberous sclerosis complex (TSC) is an autosomal dominant disorder caused by the presence of pathogenic germline variants in *TSC1* or *TSC2* genes. However, most *TSC1* and *TSC2* genetic testing strategies are limited and about 20% of patients have inconclusive or negative results. In this study, we standardized a low-cost technique based on RT-PCR followed by Sanger sequencing, aimed to complement genetic testing strategies. Patients with clinical suspicion of TSC and *TSC1* and *TSC2* inconclusive (n=3) or negative (n=1) genetic testing results were recruited. Exon 5 skipping in *TSC1* gene was identified in one patient with a previous negative genetic testing result. In addition, a 9-base-pair intronic inclusion was found in the *TSC1* of an individual with a previously identified variant of uncertain significance, c.664-10A>C. No alterations were found in the other two patients. The present study was successful in identifying errors in mRNA processing in two TSC patients with previously negative or inconclusive genetic testing results. Furthermore, this strategy was useful to reclassify a VUS to pathogenic. Finally, the validated technique could be used to complement coding region genetic tests and to investigate causality of VUS in TSC and other monogenic diseases.

**Highlights:**

- A novel cDNA analysis method revealed the presence of exon 5 skipping in the
- *TSC1* gene
- A nine-base-pair retention in TSC1 mRNA is caused by the intronic variant c.664-10A>C
- The variant of uncertain significance c.664-10A>C in *TSC1* was reclassified as pathogenic
- A strategy using cDNA could be a good complement to conventional molecular diagnosis protocols

## 1. Background

Tuberous Sclerosis Complex (TSC) is a genetic and hereditary disorder with an estimated prevalence of 1:6000 to 1:10000 **[1]**. TSC is characterized by a several disorders, including abnormalities of the skin (hypomelanotic macules, confetti lesions, facial angiofibromas, shagreen patches among others), brain (cortical tubers, subependymal nodules and giant cell astrocytomas [SEGAs], seizures, cognitive deficiencies and developmental delay, psychiatric illnesses), kidneys (angiomyolipomas, cysts, renal cell carcinomas), heart (rhabdomyomas, arrhythmias) and lungs (lymphangioleiomyomatosis [LAM] and multifocal hyperplasia) **[2-4]**. Despite the existence of specific clinical diagnostic criteria for TSC, the definitive clinical diagnosis is not always straightforward due to variable expressivity.

Therefore, the identification of a pathogenic germline variant in *TSC1* (OMIM: 191100) or *TSC2* (OMIM: 613254) allows the ultimate TSC diagnosis **[2]**. In general, 80-85% of TSC suspected cases have a detectable germline pathogenic variant in *TSC1* and *TSC2* **[1]**. The remaining 15-20% of suspected TSC patients with no mutation identified (NMI), may be explained by sampling issues (especially in mosaic individuals) **[5-6]**, deep intronic pathogenic variants (since most diagnostic tests only assess coding and canonical splice site regions) **[7-8]** and other alterations that could result in mRNA processing errors. Such errors have been previously reported in *TSC1* and *TSC2* **[9-11]**, as well as in other disease-related genes **[12-15]**. Another issue faced in the genetic testing is the lack of evidence for interpretation and classification of variants. In this scenario, variants of uncertain significance (VUS) and/or false negative variants could often be reported, which generates inconclusive results and complicates patient follow up and counseling, causing additional anxiety in the family [16]. Although additional molecular strategies such as whole exome sequencing **[17]** or RNA/coding DNA sequencing **[9**,**11]** may be useful to detect deep intronic variants or messengerRNA (mRNA) splicing errors, they are not always accessible to patients especially in underserved areas of developing countries.

In this context, an alternative low-cost technique to investigate mRNA processing defects caused by novel intronic variants or VUS is reverse transcription (RT-PCR) analysis. This technique has already been used for functional VUS studies in other diseases, such as mucopolysaccharidosis type VI **[18]**, inherited retinal diseases **[19]**, breast cancer **[20**,**21]**, and others **[22-24]**. The present study aimed to standardize a low-cost approach to analyze possible mRNA processing errors in patients with clinical suspicion of TSC and inconclusive or negative genetic testing results.

## 2. Methodology

### 2.1 Patients and ethical aspects

This is an experimental cohort study with a convenience sampling strategy. All patients that were being followed at the Oncogenetics Clinics at Hospital de Clínicas de Porto Alegre, a tertiary care center in Southern Brazil, and who fulfilled the following inclusion criteria were invited to participate in the study: suspected TSC according to clinical diagnostic criteria **[1]** and previous negative or VUS genetic testing results. In total, seven patients were contacted, and four patients were recruited (one with a previous negative and three with VUS genetic testing results). The recruitment period was between August 2022 to January 2023. All patients were born in the state of Rio Grande do Sul, Brazil. Patient’s clinical data were collected from medical charts by a medical geneticist during the recruitment period. Previous genetic testing had been conducted in 2016 by our group using a combination of a next-generation sequencing (NGS) of *TSC1* and *TSC2* coding sequences and 25 base pair (bp) exon padding combined with multiplex ligation-dependent probe amplification (MLPA) technique **[25]**. This study was approved by the Research Ethics Committee of Hospital de Clínicas de Porto Alegre under the protocol number 44739715.7.0000.5327. All patients signed an informed consent form to participate in this study. In addition to the four TSC probands, we recruited five relatives of the patient carrying a VUS to evaluate variant segregation in the family. All relatives signed an informed consent to participate in the study.

### 2.2 Samples and cell culture

Peripheral blood samples were collected in two 4mL EDTA tubes from each patient and relatives. Both tubes from each individual were processed using Ficoll-paque (*Cytiva* - GE17-1440-02) for the separation and collection of peripheral blood mononuclear cells (PBMC). Total blood (4mL) was diluted with an equal amount of PBS 1X in a 15 ml conical tube. This solution was carefully transferred to a new 15ml conical tube that had been previously filled with 4 ml of Ficoll-paque. The tube was subsequently centrifuged at 454g for 30 minutes, with the centrifuge brake off. After centrifugation, approximately 1.5ml of PBMCs were collected and transferred to another 15 ml conical tube. The PBMCs were then washed twice using 4 ml PBS 1X and spin down at 450g for 10 minutes. Finally, the cells were resuspended in 4 ml of the RPMI-1640 (Gibco - 61870036) supplemented with 20% Fetal Bovine Serum FBS (Sigma-Aldrich - F2442), 1% penicillin/streptomycin (Invitrogen - 15140148), and 1.5% Phytohemagglutinin-M (PHA-M) (Sigma-Aldrich - L1668).

PBMCs isolated from the first EDTA tube were immediately used for total RNA extraction using the PureLink RNA Mini Kit (Invitrogen - 12183020) followed by conversion to coding DNA (cDNA) using High-Capacity RNA-to-cDNA Kit (Applied Biosystems - 4387406), according to manufacturer’s instructions. PBMCs from the second EDTA tube were established in short-term cell cultures using RPMI-1640 (Gibco - 61870036) supplemented with 20% FBS, 1% penicillin/streptomycin and 1.5% PHA-M (Sigma-Aldrich - L1668) in T-25 culture flasks incubated at 37ºC and 5% CO_2_ for three days. In the third day of cell culture, cells were treated with 200μg/ml of dissolved puromycin and incubated for four hours in 5% CO_2_ and 37ºC in order to inhibit the nonsense-mediated decay (NMD) pathway, as previously described by Messiaen & Wimmer (2012) **[26]**. After that, the total volume of cells in each T-25 flask was collected and transferred to 15 ml conical tubes. The cells were spin-down by centrifugation at 450g for 10 minutes and resuspended in RPMI medium with 20% FBS, 1% penicillin/streptomycin and 1.5% PHA-M. Total RNA was immediately extracted from the cultured cells and converted to cDNA using the aforementioned commercial kits.

### 2.3 cDNA analysis

Specific primer pairs were designed to cover all *TSC1* (NM_000368.5) and *TSC2* (NM_000548.5) exons in blocks, which resulted in seven and 12 primer blocks for *TSC1* and *TSC2*, respectively. RNAm sequences available at the National Center for Biological Information (NCBI) database were used as reference sequences for primer design. Primer sequences and exon coverage of each block are described in Supplementary Information - **Table S1**. Primers were designed at the exon-exon junctions to avoid eventual DNA amplification, since DNA could be present in the initial extracted RNA samples. The cDNA targets of patients and relatives were subsequently amplified by PCR. See Supplementary Information Methods for details. Specific conditions for each primer block reaction is available in **Table S1**.

A commercial cDNA of human normal tissues from major organs of five donors (BioChain - C4234565-R) was used as reaction control to standardize each primer amplification. After the PCR reaction, all amplicons were visualized in 2% agarose gel electrophoresis using 1ul of FluoSafe dye (Alostérica - ALO35867-500) and 3ul of each PCR amplicon. A molecular ladder (3ul, Invitrogen - 10416014) with 1ul of FluoSafe dye was included in each electrophoresis to analyze the size of each amplicon. Amplicons with altered size were purified using the ExoSAP-it kit (*Applied Biosystems -* 78205) in a Proflex thermal cycler (AppliedBiosystems, Waltham, Massachusetts, EUA). The final volume of each purifying reaction was 7µl, consisting of 2µl of the ExoSAP enzymes and 5µl of PCR amplicon. Thermal cycler conditions were: 37ºC for 45 minutes to clean-up the reaction, followed by a stage at 80ºC for 15 minutes to inactivate the enzymes. Subsequently, 3ul of the purified amplicons, 2.5ul of H_2_0 and 0.5ul of specific primer previously diluted to 0.5uM/ul were added in a 0,2ml tube to be sequenced by the Sanger sequencing technique to confirm and identify the alteration.

Sanger sequencing was performed using the ABI 3500 sequencer (Applied Biosystems, California, USA), and gene sequences were compared with reference cDNA sequences from the NCBI database (NM_000368.5 for *TSC1* and NM_000548.5 for *TSC2*), through alignment using the CodonCode Aligner program. The general methodology is summarized in a flowchart in **Figure 1**. Raw data for Sanger sequencing are available in supplementary material.

**Figure 1:**
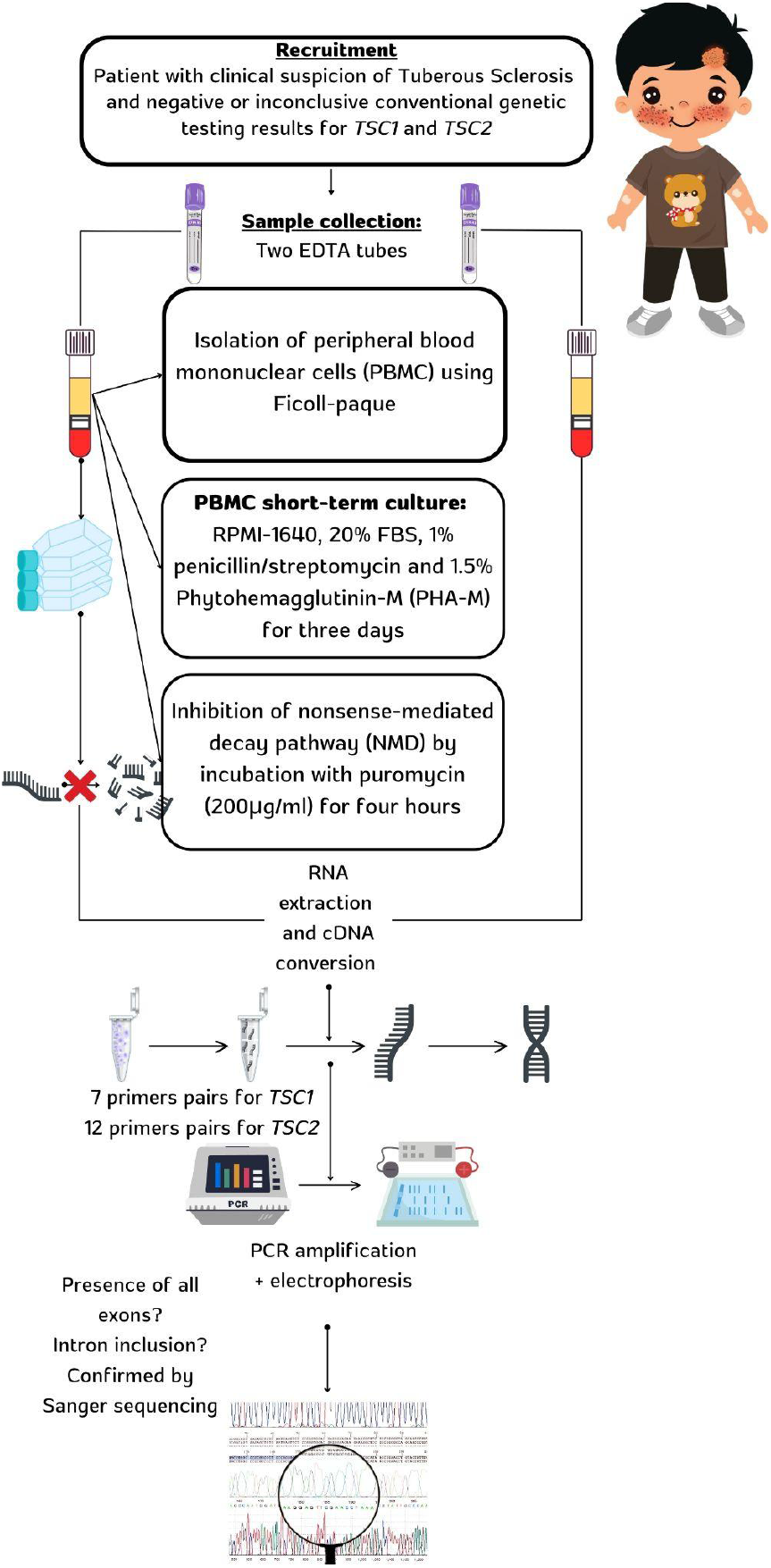
Flowchart of the methodology used in the study.

## 3. Results

A summary of the demographic, clinical and molecular features of the patients included in the study is shown in **Table 1**.

**Table 1.** Clinical and molecular features of the individuals included in this study.

| Patient | Sex/age range at testing | Genetic testing result* | Region evaluated** | Primer block used*** | Clinical manifestations |
| --- | --- | --- | --- | --- | --- |
| 1 | F/55-60 | c.975+8 G>A (*TSC2*) | intron 10 | TSC2_3<br>TSC2_4 | Facial angiofibroma, hypomelanotic macules, hepatic and renal angiomyolipoma, ungual fibromas |
| 2 | F/25-30 | Negative | All exons and introns | All primer blocks | Hypomelanotic macules, facial angiofibroma, Shagreen patches, cortical tubers, retinal astrocytoma |
| 3 | F/25-30 | c.976-3C>G (*TSC2*) | intron 10 | TSC2_3<br>TSC2_4 | Hypomelanotic macules, facial angiofibroma, subependymal nodules |
| 4 | M/15-20 | c.664-10A>C (*TSC1*) | intron 7 | TSC1_3 | Hypomelanotic macules, subependymal nodules, cortical dysplasia, Cortical renal cysts |
| Relatives of Patient 4 included in this study |  |  |  |  |  |
| Relatives of patient 4 | Sex/age range at testing | Genetic testing result* | Region analyzed** | Primer block used*** | Clinical manifestations |
| Father | M/40-45 | c.664-10A>C (*TSC1*) | intron 7 | TSC1_3 | Cortical renal cysts; Subependymal giant cell astrocytoma (SEGA) and cortical tubers |
| Mother | F/35-40 | negative | intron 7 | TSC1_3 | No clinical manifestation |
| Paternal Grandparents | - | negative | intron 7 | TSC1_3 | No clinical manifestation |
| Brother | M/1-6 | negative | intron 7 | TSC1_3 | No clinical manifestation |

M= male, F=female
\* VUS statuses were revised in the ClinVar database in December, 2023.\*\* Sequence transcript references for *TSC1* and *TSC2* genes are NM\_000368.5 and NM\_000548.5, respectively.
\*\*\* Information on the primer blocks used in this study is in Table S1.

At first, the analysis of patient 1, with the variant in *TSC2* c.975+8 G>A, and patient 3, with the variant in *TSC2* c.976-3C>G, was performed using the primer pairs TSC2_3 and TSC2_4, which covers the regions that could be affected by a variant in *TSC2* intron 10. The amplification of these regions did not show a potential exon skipping or intron retention in the cDNA samples of both patients, and thus did not contribute to clarify the role of these variants in the corresponding protein.

### 3.1 TSC2 exon 26 and 32 absence: identification of two alternative isoforms

In the analysis of patient 2, who had a negative genetic testing result, all primer pairs were amplified and analyzed. Two regions of the TSC2 cDNA were found to be altered in electrophoresis analysis: exons 23-27 (primer block TSC2_7) and exons 30-33 (primer block TSC2_9). Sanger sequencing analysis revealed the lack of exon 26 and of the first codon (CAG) of exon 27, as shown in Figure 2 (panels a and b). Furthermore, the absence of exon 32 (Figure 2c-2d) was also detected. These alterations were also found in the commercial cDNA sample, suggesting that it is not related to disease. Then, we amplified both amplicons in the additional TSC patients. The alterations were identified in all the analyzed individuals, as shown in Figure 2.

**Figure 2:**
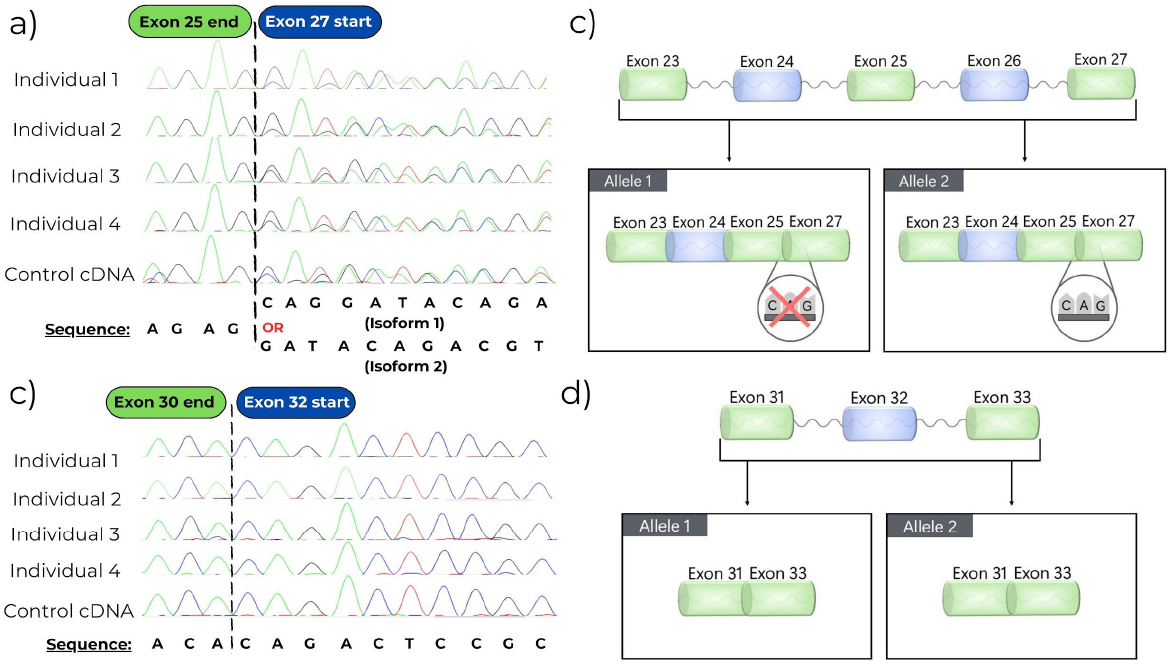
Two alternative isoforms in *TSC2*. a) Sanger sequencing of *TSC2* exons 23-27 showing absence of exon 26 in homozygosity and absence of the first codon of exon 27 (CAG) in heterozygosity in four patients with clinical suspicion of TSC and in control cDNA; b) illustrative image of the absence of exon 26 and first codon of exon 27 in *TSC2* in heterozygosity; c) Sanger sequencing of *TSC2* exons 30-33 showing the absence of exon 32 in homozygosity in four patients with suspected TSC and in control cDNA; d) illustrative image of the absence of *TSC2* exon 32 in homozygosity.

### 3.2 TSC1 exon 5 skipping

As previously mentioned, patient 2, who had no variant detected in previous genetic testing of *TSC1* and *TSC2* coding regions and intron-exon boundaries, was analyzed by amplification of all primer blocks. Three primer pairs (primer block TSC2_7, TSC2_5 and TSC2_8) did not result in reliable PCR results, and thus were not analyzed. Unfortunately, we were not able to design novel primer pairs for these regions **(Table S1)**. Amplification of the *TSC1* region covered by primer the block TSC1_2 (exons 4 to 6) showed an exon 5 skipping. cDNA from all other patients with TSC and a control cDNA were amplified and sequenced to verify if this skipping only occurred in patient 2 **(Figure 3)**. The other three patients and control cDNA did not present exon 5 skipping.

**Figure 3:**
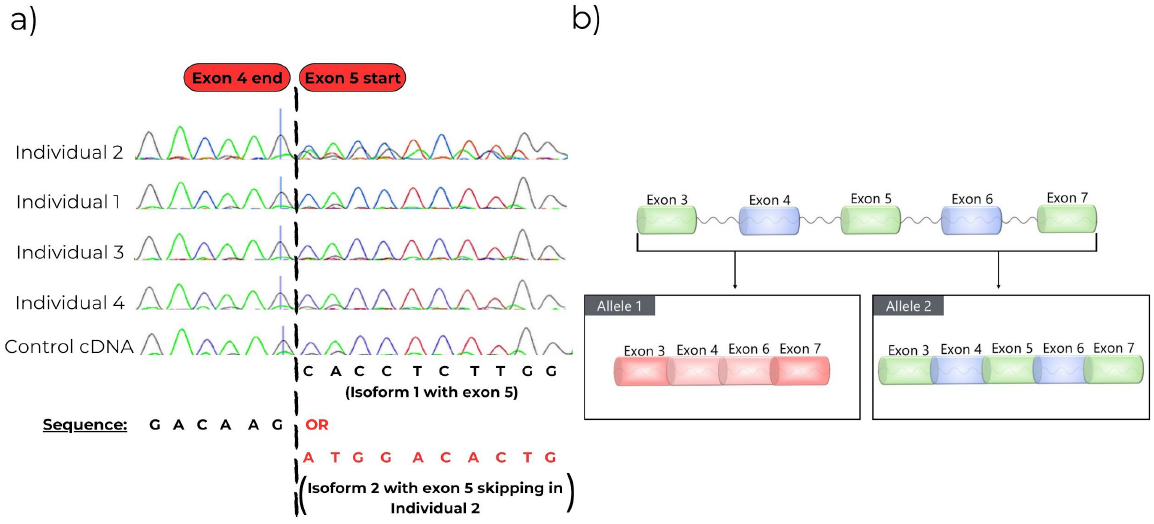
Exon 5 skipping in the *TSC1* gene. a) Sanger sequencing of *TSC1* exons 3 to 7 showing exon 5 skipping in heterozygosity in patient 2, with suspected TSC. The other three patients and control cDNA did not present exon 5 skipping; b) illustrative image of *TSC1* exon 5 skipping in heterozygosity, showing allele 1 (altered) and allele 2 (without exon 5 skipping).

### 3.3 TSC1 Intronic inclusion and Segregation analysis

To study the role of *TSC1* c.664-10A>C in the acceptor splice site in intron 7, the region spanning exons 6-10 of *TSC1* was amplified (primer block TSC1_3) using the cDNA sample of patient 4. The analysis revealed a retention of nine base pairs of intron 7 in heterozygosity, as shown in **Figure 4a-b**. Of these nine retained base pairs, the last three code for a stop codon (TAG), potentially leading to the production of a truncated protein. Then, we retrospectively analyzed the patient’s family history of TSC and genetic testing performed in his relatives. In 2018, his father, mother, paternal grandparents and younger brother were analyzed for the presence of the variant c.664-10A>C. Only his father carried the variant. The father did not report any TSC symptoms until adulthood. He currently exhibits two major and one minor TSC criteria **(Table 1)**. The clinical TSC manifestations were identified during a hospitalization to investigate headache and blurred vision that had developed over 3 months. Brain imaging showed a central nervous system tumor which, after total ventricular tumor resection and histopathology, was diagnosed as a subependymal giant cell astrocytoma (SEGA). Additionally, cortical tubers were identified. In the same year, abdominal computed tomography identified cortical renal cysts measuring 2.2 cm. In contrast to his father, patient 4 was diagnosed with hypomelanotic macules, subependymal nodules, renal cysts and cortical dysplasia in the early stages of his life. The other family members did not show any TSC clinical characteristics. The mother does not present clinical manifestations of TSC and no identifiable genetic variants in TSC1 and TSC2 are identified. Additional paternal relatives (uncle and aunt) do not manifest any TSC symptoms, but they were unavailable for genetic testing.The family pedigree is shown in **Figure 4c**.

**Figure 4:**
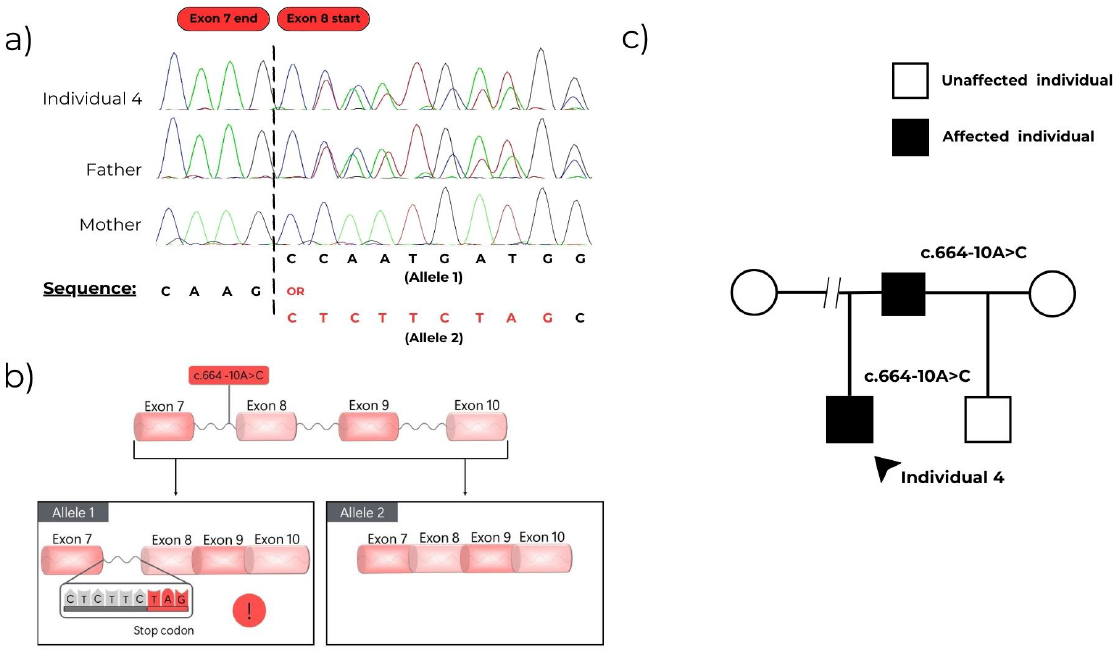
*TSC1* intronic inclusion identified in patient 4. a) Sanger sequencing of the *TSC1* region containing exons 6 to 10 showing an intronic inclusion of nine base pairs of intron 7 in the cDNA samples of patient 4 and his father; the alteration was not detected in the patient’s mother and other relatives. b) illustrative image of the intronic inclusion of nine base pairs of intron 7 in the *TSC1* transcript; c) *TSC1* c.664-10A>C segregation analysis in patient’s 4 family.

## 4. Discussion

In most patients with TSC, clinical features allow the identification of suspected cases. Approximately 85-90% of suspected patients who undergo conventional genetic testing, such as NGS with MLPA of the coding region of the *TSC1* and *TSC2* genes, have identifiable pathogenic or likely pathogenic germline variants. In the remaining cases, diagnosis is elusive **[1]**. Considering that the definitive diagnosis of TSC is made when a molecular loss of function alteration is identified in either *TSC1* or *TSC2* genes, other less common alterations resulting from mRNA processing errors should be sought when routine genetic testing is reportedly inconclusive. Some costly and high-throughput techniques can be employed to resolve these cases, such as RNA-seq. However, the current study aimed to employ a low-cost analysis. The average cost of this analysis in Brazil for VUS carriers and negative cases was approximately $46.26 and $440 per sample, respectively. In contrast, RNA-seq average cost is approximately $1000 per sample. Previous studies have described mRNA processing errors leading to aberrant isoforms and TSC phenotype using different methodologies. Notably, Tyburczy *et al*. (2015) **[10]** described the *TSC1* c.664-15A>G variant, located in the intron 7 acceptor splice site, resulting in a premature termination codon and the *TSC2* c.976-15G>A variant in intron 11, resulting in a frameshift and loss of function of the gene.

In the present study, we developed and validated a laboratory protocol to identify mRNA processing errors and assessed a series of patients with inconclusive germline testing results (either negative or VUS). VUS statuses were reviewed in ClinVar by December, 2023 **[27]**. One of them (c.975+8 G>A) had novel multiple submissions as likely benign and benign. However, there were no functional and segregation studies to confirm the status of this variant, and therefore we decided to include it in the present study. The c.976-3C>G variant has been submitted to the ClinVar database with no functional evidence since September, 2013 **[27]**. The LOVD database showed three submissions of this variant as pathogenic, with supporting evidence indicating the presence of aberrantly spliced transcripts. The variant has been reported in the literature as IVS9-3C>G in two studies published in 1999 and 2000, which are associated with two aberrant transcripts: exon 11 skipping and a novel cryptic splice acceptor in exon 11, which results in the lack of the first 56 bp of exon 11[**28, 29]**. However, we were not able to replicate this finding using our strategy.

In the analysis of the four TSC patients recruited in this study, we identified two alterations that were present in all of them. The first alteration, *TSC2* exon 26 absence, was previously described by Xu *et al*. (1995) **[30]**, who analyzed cDNA samples from human adult and fetal tissues, as well as rat tissues, and identified normal tissues with and without *TSC2* exon 26. Interestingly, they showed that in many human adult tissues express both isoforms (with and without exon 26), but lymphoblasts and fibroblasts completely lack exon 26. In addition, they showed that the first codon of *TSC2* exon 27 could be absent in human lymphoblasts and fibroblasts, usually in heterozygosity. The first codon of exon 27 was also found to be absent in individuals from China without clinical manifestations of TSC **[31]**. In our study using peripheral leukocyte samples, all patients displayed the absence of exon 26 in homozygosity and the absence of the first codon of exon 27 in heterozygosity. The second alteration found in all the recruited TSC patients was *TSC2* exon 32 absence. The presence of *TSC2* exon 32 has been reported in human fetal brain, kidney, and heart tissues. However, mRNA expression without exon 32 seems to be also common in the human fetal brain and kidney **[30-31]**. Conservation analysis of exons 26 and 32 has shown that they are not highly conserved and that generally their absence is unrelated to TSC **[32]**. Interestingly, the same authors reported one specific variant in exon 32 that seems to be associated with epilepsy, but it is not related to TSC (c.3846_3855delinsG).

The finding of a potential loss of function *TSC1* exon 5 skipping in patient 2 is also significant and probably results from a deep intronic variant in introns 4 or 5. We have a perspective to investigate these intronic regions. To date, pathogenic variants in the same intron have been reported in both *TSC1* and *TSC2* which cause mRNA processing errors **[9-11]**. The N-terminal region, which encompasses exon 5 of the *TSC1* gene, does not have a functional domain with a known established function, but certain pathogenic variants in this regions could destabilize the TSC complex, and cause loss of function in the regulation of the mTOR pathway **[34-36]**. Furthermore, skipping of *TSC1* exon 5 due to the exonic variant c.362A>G (p.Lys121Arg) has been identified in a patient with *de novo* TSC **[37]**.

Finally, we identified that the *TSC1* c.664-10A>C variant in patient 4 changes a splice site by inserting nine base pairs at the end of intron 7 in mRNA (CTCTTCTAG). This insertion creates three new codons in the corresponding protein, leucine (Leu), phenylalanine (Phe), and a termination codon (TAG) - which results in a premature termination (PTC) and production of a truncated hamartin. It is expected that this variant results in a hamartin with only 223 amino acids, where 221 are encoded up to exon 7 and the remaining two result from intronic retention. The truncated protein lacks the coiled-coil domain, its main functional region, and the region that interacts with tuberin, encoded by the *TSC2* gene **[38, 39]**. Since the mRNA with PTC could be targeted to the nonsense-mediated mRNA decay (NMD) pathway **[40]**, we assessed NMD activity in this altered mRNA by collecting two samples: one for total RNA extraction without cell culture, and one to establish a PBMC culture treated with puromycin to inhibit the NMD pathway. Samples with and without cell culture, obtained from patient 4 and his father, both with the *TSC1* c.664-10A>C variant, exhibited the allele with intronic retention and we could not detect differences between samples with and without NMD inhibition.In this sense, Jeganathan *et al*. (2002) **[41]** found that certain transcripts carrying a pathogenic variant in *TSC1* undergo greater degradation by the NMD pathway in fibroblasts, as opposed to the lymphoid cell line. This finding could explain our results in NMD inhibited and uninhibited samples.. Notably, segregation analysis of the *TSC1* c.664-10A>C variant shows that it is only present in two patients with clinical findings of the TSC phenotype and absent in other relatives without TSC symptoms. In 2016, by the time of our previous study,, the *TSC1* c.664-10A>C variant was not reported in Clinvar and LOVD databases. However, in 2022, it was submitted to ClinVar and classified as likely benign, despite the limited evidence available regarding this variant. The intronic inclusion identified in our study resulted in an unusual stop codon, which led to the reclassification of this variant to pathogenic according to the The American College of Medical Genetics and Genomics (ACMG) criteria (BP4 + PM4 + PP2 + PM2 + PP4 + PVS1) **[42]**.

## 5. Conclusion

Tuberous sclerosis complex (TSC) is an autosomal dominant systemic disorder predisposing to multiple benign and malignant tumors in most patients. Variable clinical features allow the identification of suspected cases, but definitive diagnosis is made when a molecular loss of function variant is identified in *TSC1* or *TSC2* genes. The present study aimed to standardize a low-cost approach to analyze possible mRNA processing errors in patients with clinical suspicion of TSC and inconclusive or negative genetic testing results. This approach allowed the reclassification of the *TSC1* c.664-10A>C from VUS to pathogenic, and identified a *TSC1* exon 5 skipping in a patient with no previously identifiable alterations. We were also able to identify splicing alterations which are likely common and unrelated to disease, resulting in alternative isoforms of the corresponding transcripts. These findings reinforce the correct design of our technique. In conclusion, in suspected TSC patients with negative or inconclusive results, a low-cost approach to study mRNA processing errors may provide clarification of the diagnosis and facilitate genetic counseling of affected patients and their at-risk relatives.

## Data availability

Clinical and molecular data from this study are openly available in ClinVar at SUB15042297.

The primer sequences used to analyze the specific regions and raw data produced by Sanger sequencing of all analyzed regions are available as supplementary data.

All the data were reported in accordance with STROBE (STrengthening the Reporting of OBservational studies in Epidemiology) statements.

The manuscript was published in a preprint version in MedRxiv - MS ID#: MEDRXIV/2025/321206.

## Acknowledgments

We gratefully thank Fundo de Incentivo à Pesquisa e Eventos (FIPE) of the Hospital de Clínicas de Porto Alegre for their financial support.

## Supplementary materials

The supplementary materials contain additional information regarding the methodology, as well as a table that details the primers used in the study. Furthermore, it provides the raw data for Sanger sequencing.

- **Table S1:** Sequence and details of primer blocks used in study.
- Raw data Sanger - Patient 1
- Raw data Sanger - Patient 2
- Raw data Sanger - Patient 3
- Raw data Sanger - Patient 4
- Raw data Sanger - Control and relatives patient 4

