## Supplemental Table 1 for "*TSC1* mRNA processing alterations in tuberous sclerosis complex (TSC) patients with negative or inconclusive genetic testing results"

**Supplementary Information for article: *TSC1* mRNA processing alterations in tuberous sclerosis complex (TSC) patients with negative or inconclusive genetic testing results.**

**(Garcia *et al.*, 2025)**

This file contains supplementary information of the methodology and a table with details of primer used in the study.

### Supplementary Methods:

#### PCR reaction:

Each PCR mix (25µl total reaction) contained: 18-19.3µl ultrapure water, 2.5µl buffer with Tris-HCl 200 mM, pH 8.4, KCl - 500 mM without $MgCl_2$ (Invitrogen, 10966-030), 0.5-0.8µl dNTP - 5mM (Invitrogen - 10297018), 0.5-0.8µl $MgCl_2$ - 50mM (Invitrogen, 10966-030), 0.5µl of each primer - 5uM, 0.1µl Platinum Taq DNA polymerase - 5U/uL (Invitrogen, 10966-020), and 1ul of cDNA. PCR reactions were performed in Proflex thermal cycler (AppliedBiosystems, Waltham, Massachusetts, EUA) using the following parameters: denaturation at 94ºC for five minutes, followed by 35-40 cycles at 94ºC for 30 seconds, 54-60ºC for 30 seconds, 72ºC for 30 seconds, and a final extension step of 72ºC for two minutes.

**Supplementary table 1:** Sequence and details of primer blocks used in study.

| Primers | Forward (5'-3') | Reverse (5'-3') | Amplicon (bp) | AT* (ºC) | Cycles in PCR reaction |
|---|---|---|---|---|---|
| ***TSC1* (exons)** | | | | | |
| TSC1_1 (1-3) | GGGGAGGTGCTGTACG | CCACGGTCAGAATTGAGG | 341 | 60 | 38 |
| TSC1_2 (4-6) | GCTGGACTCCCCCATGCT | CGTGGCCTGGTTTCTTCAGG | 476 | 60 | 35 |
| TSC1_3 (7-10) | GAAGAAACCAGGCCACG | GCTCCAAAGAGTAGCTTGTG | 547 | 60 | 40 |
| TSC1_4 (11-14) | TGAACCACCACAAGCTACTC | CTAGATATTGCAGCTTCTTCTTTAT | 435 | 54 | 35 |
| TSC1_5 (15-16) | AAAGATAAAGAAGAAGCTGCAAT | AGCCTCCAAAGTGGGTC | 622 | 55.5 | 40 |
| TSC1_6 (17-19) | CTTTGGAGGCTCTCCTCCTTC | CAGCTGCCTGTTCAAG ACTC | 523 | 54 | 40 |
| TSC1_7 (20-22) | GTTTCCCAAAAGCTCTCAAAC | CCGTCATTACAACAGTCAAGC | 506 | 60 | 35 |
| ***TSC2* (exons)**** | | | | | |
| TSC2_1 (1-3) | GGGGGTGCGCCTTTCT | CCACTGCGTGCTCTTCAAAT | 320 | X | X |
| TSC2_2 (4-5) | AGAACCTCAATTCTGACCGT | GTCCATCTTGAGACATTTTAGT | 278 | 60 | 40 |
| TSC2_3 (6-10) | AGAGCTGGCTGACTTTGTCCT | CATGGCCTGGTAAAATGATGGC | 508 | 60 | 35 |
| TSC2_4 (11-15) | GTCCGAACGAGGTGGTGTC | GGGCCATACCTTCTCGATG | 624 | 55 | 35 |
| TSC2_5 (16-19) | CTGCCACACACACCACTTCAA | TTCCAGTCAGACTCCTGCTTCAA | 558 | X | X |
| TSC2_6 (20-22) | TGCCCTACTCCCTGCTCTT | GCCAGAGTGGACAGGAACTC | 502 | 60 | 35 |
| TSC2_7 (23-27) | TGGAGTTCCTGTCCACTC | CAGGAGACCTCTTCGGG | 607 | 58 | 35 |
| TSC2_8 (28-30) | GGAGCTCACGGAAACCTGTC | GCTGGTGTTCCCTGTGGG | 562 | X | X |
| TSC2_9 (31-33) | GAGGCCCACAGGGAAC | CTGGAGACTGAGGACGAC | 423 | 59 | 35 |
| TSC2_10 (34) | GGTCGTCCTCAGTCTCC | GCAGGAACACGAAACTGG | 504 | 59 | 35 |
| TSC2_11 (35-39) | AACCCCAGTTTCGTGTTC | CTCCATGTCTTTCCTGCAC | 591 | 60 | 35 |
| TSC2_12 (40-41) | CAGGAAAGACATGGAGGG | GGCTTCCTCGCAGATCC | 214 | 60 | 40 |

***Annealing temperature of which standardized PCR reaction of primers to cover *TSC1* and *TSC2* genes.**
**** The pair primers with the word "X" in AT and cycles columns do not show reliable results in PCR reactions.**
